# Behavioral evidence of reduced working memory capacity in fibromyalgia patients is not reflected in electrophysiological outcomes

**DOI:** 10.1101/2025.09.20.25336159

**Authors:** Elizabeth L. Young, Yanina V. Atum, Christian A. Mista, Diego Arévalo, Belén Moglia, José A. Biurrun Manresa

## Abstract

Fibromyalgia is characterized by widespread musculoskeletal pain accompanied by sleep disturbances and cognitive dysfunction, among other symptoms. Patients frequently report difficulties with memory, but objective assessment of these impairments remains limited. This study aimed to evaluate working memory performance in fibromyalgia patients using two established paradigms: the change detection task, which primarily measures storage capacity, and the n-back task, which assesses both storage and manipulation of information.

For the change detection task, the behavioral outcomes assessed were the hit rate, false alarm rate, capacity estimate and response times. The electrophysiological measure evaluated was the contralateral delayed activity. For the n-back, the behavioral outcomes were the same, except for the capacity estimate. Electrophysiologically, the P2 and P3 from the evoked potentials were the outcomes of the task. Behaviorally, results demonstrated that fibromyalgia patients exhibited lower memory capacity than controls (1.90 vs 2.64), in the change detection task, which involved differences in the hit rate and false alarm rate, whereas no behavioral differences were found for the n-back task. In contrast, no differences were found for any of the electrophysiological outcomes in any of the tasks. Taken together, we found evidence supporting a reduction in working memory capacity in fibromyalgia, although it is not reflected in electrophysiological measures.

## 1. Introduction

Fibromyalgia Syndrome (FMS) is a chronic condition characterized by the presence of generalized musculoskeletal pain, along with other non-specific symptoms, such as depression, anxiety, fatigue, sleep disturbances, hyperalgesia, allodynia and cognitive impairment (Wolfe et al., 2010). The prevalence of FMS varies from one region to another, being a global estimate between 2% and 8%. Additionally, the prevalence is larger in women than men with a proportion of 4:1 (Cabo-Meseguer et al., 2017; Dizner-Golab et al., 2023). While generalized musculoskeletal pain is the predominant symptom, cognitive impairment has also been reported (Bell et al., 2018; Glass, 2009). The term “fibrofog” has been used to refer to the subjective cognitive impairment experienced by patients, alongside the term “dyscognition”, which encompass objective cognitive difficulties observed in neuropsychological tests (Dass et al., 2023; Pidal-Miranda et al., 2018). Those terms describe difficulties in concentrating, attentional and memory impairment, and inability to perform multiple tasks at once, and the prevalence of these issues is higher in fibromyalgia patients compared to other rheumatology conditions (Kravitz & Katz, 2015). Furthermore, quantitative evidence has been found of alterations in executive functions in this group of patients, predominantly related to deficiencies in inhibitory control and working memory (Bell et al., 2018).

In particular, visual working memory (VWM) refers to the domain that allows storing visual information for a short period (in the order of seconds), and its capacity has been linked to other cognitive functions (Feldmann‐Wüstefeld, 2021). Estimation of the VWM capacity has been carried out using electroencephalography (EEG) through the change detection paradigm, from which the contralateral delay activity (CDA) can be derived. The CDA is a negative slow wave observed in the EEG, and its amplitude is correlated with the number of objects maintained in the VWM (Luria et al., 2016). In a change detection task, participants are presented with a varying number of items that they have to remember. During the retention interval, the items disappear, and participants are asked to maintain them in memory. Finally, the items reappear, and participants indicate if a change has occurred (Adam et al., 2018). The capacity estimate (K) is calculated using behavioral outcomes of the change detection task and it refers to the maximum number of items that can be maintained in the VWM (Rouder et al., 2011). Studies have been carried out in pathological populations to assess changes in the CDA. Wiegand et al. (2016) found attenuation in the CDA in adult attention-deficit/hyperactivity disorder patients compared to healthy controls, indicating altered brain mechanisms underlying visual storage capacity in such pathology. In contrast, Farina et al. (2020) found that the CDA was not a robust marker for subclinical mild cognitive impairment, as previously proposed. To the best of our knowledge, no research has been done on fibromyalgia patients using the change detection paradigm. Another commonly used task to evaluate VWM capacity is the *n-back* task, which allows estimating the maintenance and manipulation of stored information, among other aspects of executive functions, such as inhibitory control (Shalchy et al., 2020). Changes in measures derived from EEG recordings, such as event-related potentials (ERPs) or power bands, allow researchers to assess memory load (Brouwer et al., 2012). In particular, ERP components related to working memory are the P2 and P3 waves. The P2 component (peaking 150-275 ms after stimulus onset) has been linked to processes of encoding information in memory, particularly in parieto-occipital scalp locations (Finnigan et al., 2011). The P3 component (peaking at around 250-600 ms after stimulus onset) has been linked to processing capacity in response to working memory tasks (Mercado et al., 2022). The behavioral outcomes of this task include hit rate, false alarm rate, number of errors and accuracy, as well as response time. Studies carried out in fibromyalgia patients have detected differences in behavioral outcomes such as response time (Tesio et al., 2015) and performance, in addition to lower amplitudes in ERPs, compared to healthy volunteers (Mercado et al., 2022). Nevertheless, there is also evidence suggesting no differences in performance between groups (Gelonch et al., 2016; Walitt et al., 2016). Considering this, we explored differences in cognitive performance in fibromyalgia patients, particularly related to VWM, by recording EEG and behavioral outcomes during change detection and n-back tasks.

## 2. Materials and Methods

### 2.1. Participants and ethical approval

Sixteen patients with clinical diagnosis of fibromyalgia and fifteen age- and gender-matched healthy volunteers participated in the study. Written informed consent was obtained from all volunteers before participation. All experimental procedures were conducted following the Declaration of Helsinki and were approved by the Central Bioethics Committee in Medical Research and Practice (Entre Ríos, Argentina), Protocol ID: IS003981. Furthermore, the protocol was preregistered in ClinicalTrials.gov (NCT number: NCT05910372).

### 2.2. Sample size estimation

Sample size was estimated using the hit rate in the change detection task as the main outcome. We conducted a pilot study with 10 healthy volunteers to determine the distribution of the variable across the group. Based on those results, we selected the hit rate for the highest cognitive load (i.e. 6 squares to be remembered) as the measure with the best potential discriminative power between fibromyalgia patients and healthy controls. In the pilot study, the mean hit rate for a cognitive load of 6 squares was 75% ± 10%. We hypothesized a 20% difference between healthy volunteers (75% ± 10%) and fibromyalgia patients (55% ± 20%) considering it the minimum difference of experimental relevance (Copay et al., 2007). Taken together, the sample size required to achieve a significance level of 5% and a statistical power of 80% on a two-tail independent sample t-test was 14 participants per group.

### 2.3. Self-report instruments

Fibromyalgia patients completed the Revised Fibromyalgia Impact Questionnaire (FIQR), an instrument developed to measure both physical and psychological symptoms of fibromyalgia. The final score represents the overall impact of those symptoms on the quality of life (Bennett et al., 2009; Salgueiro et al., 2013). Healthy volunteers answered the equivalent questionnaire, i.e., the Symptoms Impact Questionnaire (SIQ). The Hospital Anxiety and Depression Scale (HADS) was used to assess the levels of anxiety and depression, we used (Vallejo et al., 2012). Finally, the Short Form of the Brief Pain Inventory (SF-BPI) was used to measure multiple aspects of pain, such as intensity (i.e. worse, least, average, current) and functional interference of pain in daily activities. It also assesses the pain medication use, as well as the percentage of relief (Williams & Arnold, 2011). All questionnaires were implemented in their validated Spanish versions.

### 2.4. Cognitive tasks

Participants underwent a single experimental session consisting of 10 blocks of 30 trials each of the change detection task and 5 blocks of 62 trials each of the *n-back* task, with a short break (1-2 min) between blocks. Behavioral outcomes derived from the participants’ response were assessed while they were executing the tasks, concurrently with EEG recordings.

#### 2.4.1. Change Detection Task

Participants were instructed to fixate on a black dot at the center of a computer screen, present at all times, to avoid eye movement artifacts in the EEG recordings. Each trial began with a blank screen (500 ms) followed by a green arrow (1100 ms) indicating the side of the display to be remembered. Then, the memory array, i.e., the sets of an equal number of colored squares on the cued and uncued sides, was presented for 250 ms. This interval is known as the *encoding phase*. The set sizes to be memorized were 2, 4, and 6 squares. The *retention period* followed the encoding phase and consisted of another blank screen with a duration of 1300 ms. After the retention period, participants were presented with a new set of colored squares, and they had to answer whether the squares were the same color and at the same location as those presented in the encoding phase or not. The response was self-paced and was recorded by clicking left (“same set”) or right (“different set”) on a computer mouse (Figure 1, top). A block of this task contained 10 trials of each set size, totaling 30 trials per block.

**Figure 1:**
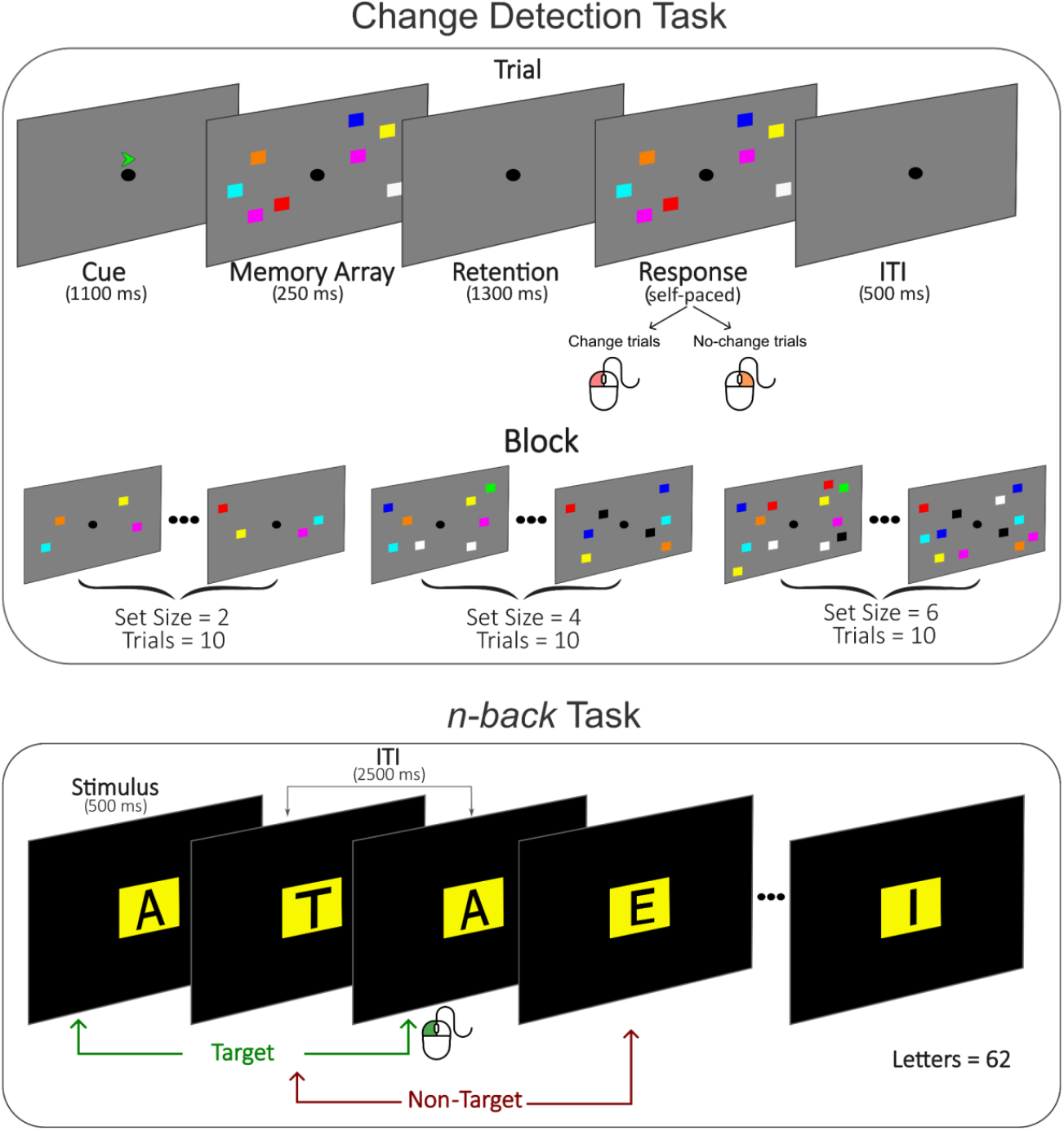
Detail of the experimental design. (Top) Change detection task. (Bottom) n-back task

#### 2.4.2. N-back task

Participants performed a *two-back* memory task, in which they were instructed to continuously observe the center of the display, as a sequence of 62 letters appeared one at a time. The appearance of the letter marked the beginning of each trial and remained on the screen for 500 ms. The task consisted of detecting whether the current letter on the screen was the same as the one presented two trials prior in the sequence (*target stimuli*). The inter-trial interval was 2500 ms, and within this time window, participants were instructed to provide a response if a target stimulus was detected using a computer mouse. Participants were instructed not to respond to non-target stimuli. The percentage of target stimuli was set to 33% (Figure 1, bottom).

### 2.5. Behavioral measures

For the change detection task, the hit rate (*H*) was derived as the proportion of correct change trials, whereas the false alarm rate (*F*) was calculated as the proportion of incorrect no-change trials. With that, VWM capacity (*K*) was estimated as *K*= *N(H−FA)*,with N representing the set size (Cowan, 2001). For the *n-back* task, the behavioral outcomes were the hit rate and the false alarm rate. Additionally, response times were recorded for both tasks.

### 2.6. EEG

#### 2.6.1. Recording

Continuous EEG data was recorded with a Neuroscan SynAmps amplifier (Compumedics, Australia), using 15 electrodes placed according to the 10-20 International System. Ground and reference electrodes were placed on the right and left ear lobes, respectively. Data was recorded with a band-pass filter (0.15-30 Hz) and sampled at 250 Hz.

#### 2.6.2. Pre-processing

EEG data was analyzed offline using Python’s MNE (Gramfort, 2013; Larson et al., n.d.) library. For each participant, continuous data was low-pass filtered at 30 Hz using a Butterworth IIR digital filter. Afterwards, independent component analysis (ICA) was applied to perform artifact rejection by manually selecting components related to blinking and muscular artifacts.

#### 2.6.3. Contralateral delay activity processing

To obtain the CDA waveform, the pre-processed data was divided into epochs from 600 ms pre-stimulus to 1750 ms post-stimulus, with no baseline correction. Only correct trials were used in the subsequent analysis. Lateralized waveforms were computed by subtracting the average of the ipsilateral electrodes from the average of the contralateral electrodes for each set size and each volunteer (Adam et al., 2018; Luria et al., 2016). The selected electrodes for the analysis were PO3/PO4 and PO7/PO8.

#### 2.6.4. N-back processing

The pre-processed data recorded when performing the *n-back* task was divided into epochs from 800 ms pre-stimulus to 1700 ms post-stimulus, with no baseline correction. Correct target and non-target responses were selected and averaged for each participant. The grand-average waveform was computed by averaging across participants for each condition.

### 2.7. Statistical analysis

Descriptive statistics are presented as mean ± standard deviation or median [interquartile range], depending on the underlying probability distribution of each variable. The statistical analysis of the behavioral data was carried out in Jamovi v.2.3.28. For the change detection task, differences in hit rate, false alarm rate and capacity were assessed using a Welch’s independent sample t-test with *condition* (fibromyalgia patient or healthy volunteers) as the grouping factor. The statistical analysis for the response times was performed by fitting a generalized mixed model, with *condition* (fibromyalgia patients or healthy volunteers) as a fixed factor, *set size* (2, 4, or 6) as a covariate, and random intercept for *subjects*. The response variable was modelled with a gamma distribution and a log link function. For the *n-back* task, differences in hit rate, false alarm rate, and response times were assessed using a non-parametric Mann-Whitney test. Differences in CDA amplitudes between conditions for each set size and differences in amplitudes of the waveforms derived from the *n-back* task were analyzed using a non-parametric cluster-level permutation test, using the Python package MNE. P values smaller than 0.05 were regarded as statistically significant, but results were further interpreted considering effect sizes and minimal experimentally relevant differences as well (Mista et al., 2023; Schober et al., 2018; Sullivan & Feinn, 2012).

## 3. Results

### 3.1. Demographic and clinical data

.Demographic and clinical data are summarized in Table 1.

**Table 1:** Demographic and clinical results

| Variables |  | Fibromyalgia Patients (n=16) | Healthy participants (n = 15) |
| --- | --- | --- | --- |
| Age (years) |  | 44.9 ± 9.1 | 45.9 ± 9.8 |
| Gender |  | Female (16) | Female (15) |
| Time from diagnosis (years) |  | 6.5 [6] | - |
| Maximum educational level completed | Elementary School | 1 | 0 |
|  | Highschool | 9 | 3 |
|  | Undergraduate | 6 | 6 |
|  | Graduate | 0 | 6 |

Results of the self-report instruments show a total score of 69.3 [20] and 10.3 [9.6] for the FIQR and SIQR, respectively. A Mann-Whitney U test showed statistically significant differences (U = 10, p < 0.001) between the scores of each group. Anxiety scores derived from the HADS questionnaire showed scores of 14.5 [6] for fibromyalgia patients and 5 [2.5] for control, resulting in statistically significant differences (U =15, p < 0.001). Depression scores, also obtained from the HADS questionnaire, are 11 [4.5] for patients and 1 [1.5] for healthy volunteers, and statistically significant differences between both groups (U = 12.5, p < 0.001). From the BPI, we analyzed only the pain scores at the time of the experiment, resulting in a score of 6 [2.25] for patients and 0 [0] for healthy volunteers. From statistical analysis, significant differences were found (U = 10.5, p < 0.001).

### 3.2. Behavioral measures

#### 3.2.1. Change detection task

Figure 2 and Figure 3 shows the hit rate and the false alarm rate of both groups for the different set sizes. For fibromyalgia patients, the hit rate was 0.89 [0.07] for set size 2, 0.72 [0.18] for set size 4 and 0.58 [0.28] for set size 6. For healthy volunteers, it was 0.9 [0.07] for set size 2, 0.8 [0.15] for set size 4 and 0.69 [0.21] for set size 6. Upon statistical analysis, we found significant differences between groups for set sizes 4 and 6 (*t*_*24*_ = *3*.*107, p* = *0*.*002* and *t*_*25*.*2*_ = *2*.*486, p* = *0*.*01*, respectively), but not for set size 2 (*t*_*20*.*3*_ = *0*.*981, p* = *0*.*169*). The false alarm rate for fibromyalgia patients was 0.04 [0.13] for set size 2, 0.12 [0.13] for set size 4 and 0.1 [0.14] for set size 6. For healthy volunteers it was 0.02 [0.05] for set size 2, 0.08 [0.07] for set size 4 and 0.13 [0.09] for set size 6. The statistical analysis revealed significant differences between groups for set sizes 2 and 4 (*t*_*16*.*2*_ = −*2*.*01, p* = *0*.*031* and *t*_*19*.*4*_ = −*2*.*3, p* = *0*.*017*), but not for set size 6 (*t*_*21*.*6*_ = −*1*.*06, p* = *0*.*150*).

**Figure 2:**
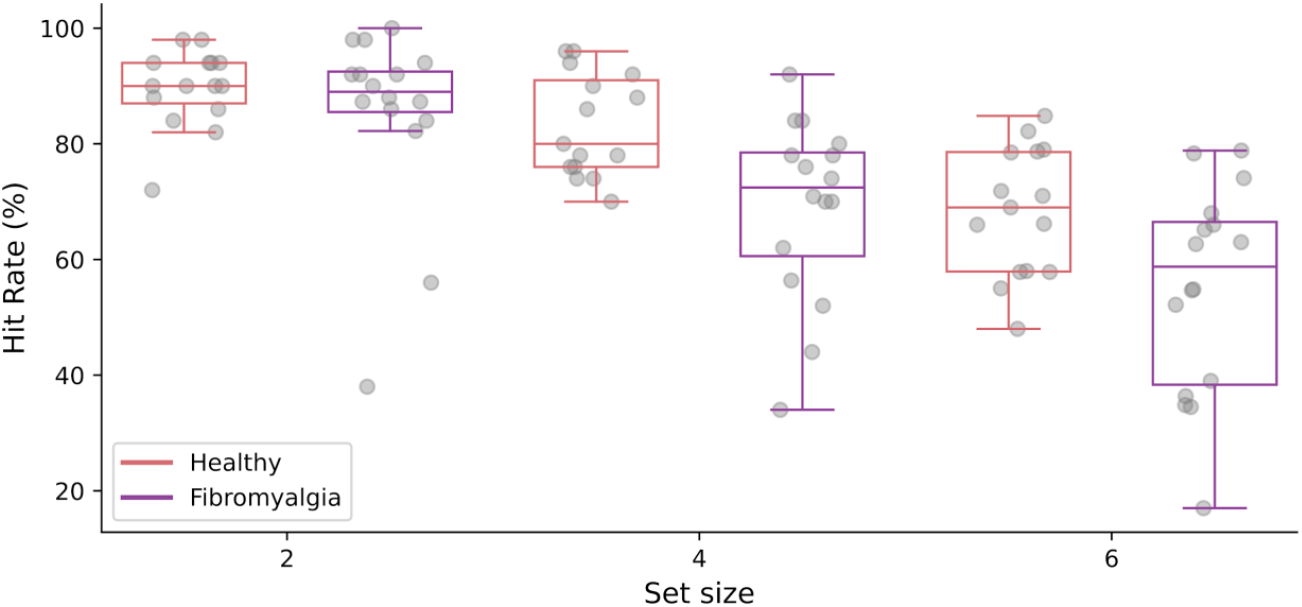
Hit rate for fibromyalgia patients and healthy volunteers in the Change Detection Task, for each set size

**Figure 3:**
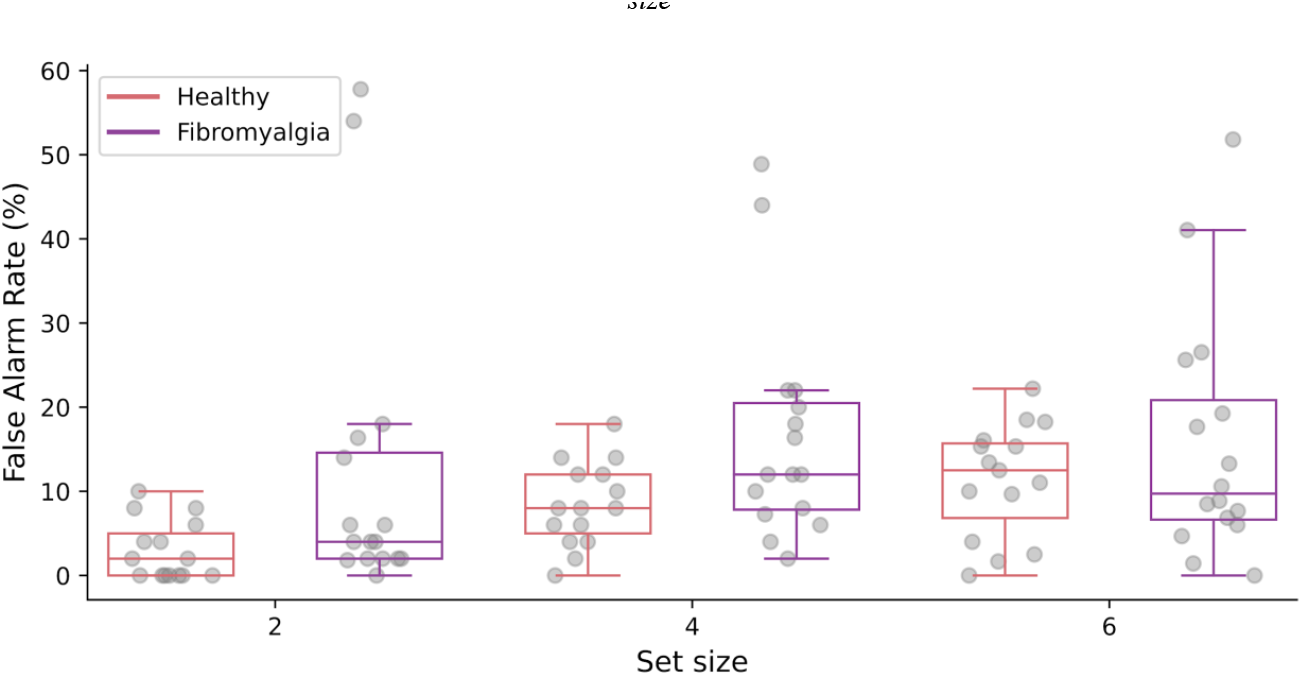
False alarm rate for fibromyalgia patients and healthy volunteers in the Change Detection Task, for each set size

Figure 4 (left) shows the VWM capacity estimates for participants of both groups and for each set size. For fibromyalgia patients, K was 1.73 [0.28] for set size 2, 2.48 [1.12] for set size 4 and 2.56 [1.53] for set size 6. For healthy volunteers, K was 1.8 [0.18] for set size 2, 2.8 [0.8] for set size 4 and 3.39 [1.09] for set size 6. Statistical analysis shows significant differences between groups for set sizes 4 and 6 (*t*_*23*.*2*_ = *3*.*379, p* = *0*.*001*, for set size 4;*t*_*25*.*6*_ = *2*.*690, p* = *0*.*006*, for set size 6). The average capacity was 1.90 [1.08] for fibromyalgia patients and 2.64 [1.55] for healthy volunteers (Figure 4, right). Statistical analysis revealed significant differences between the two groups (*t*_*90*.*6*_ = *3*.*65, p* < *0*.*001*). Statistical analysis of hit rate and false alarms for the change detection task are provided in the Supplementary Section.

**Figure 4:**
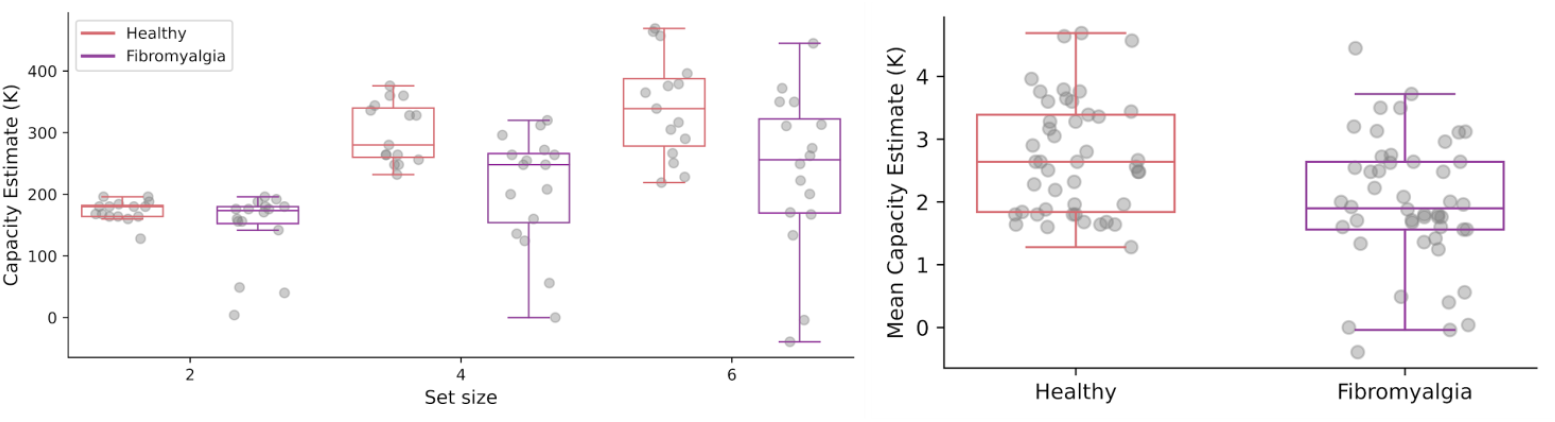
(Left) Capacity estimate for fibromyalgia patients and healthy volunteers in the Change Detection Task, for each set size. (Right) Average capacity estimate

Figure 5 shows the distribution of response times for both groups and each set size. For fibromyalgia patients, response times were 1053 [368] ms for set size 2, 1304 [461] ms for set size 4, and 1448 [642] ms for set size 6. For the control group, response times were 857 [213]ms for set size 2, 955 [259] ms for set size 4, and 1054 [212] ms for set size 6. To quantitatively assess response times for patients and controls for the different set sizes, we fitted a generalized mixed model. We found a significant main effect for set size 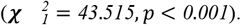 We did not find a significant main effect for condition 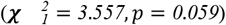 nor interactions 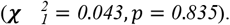

**Figure 5:**
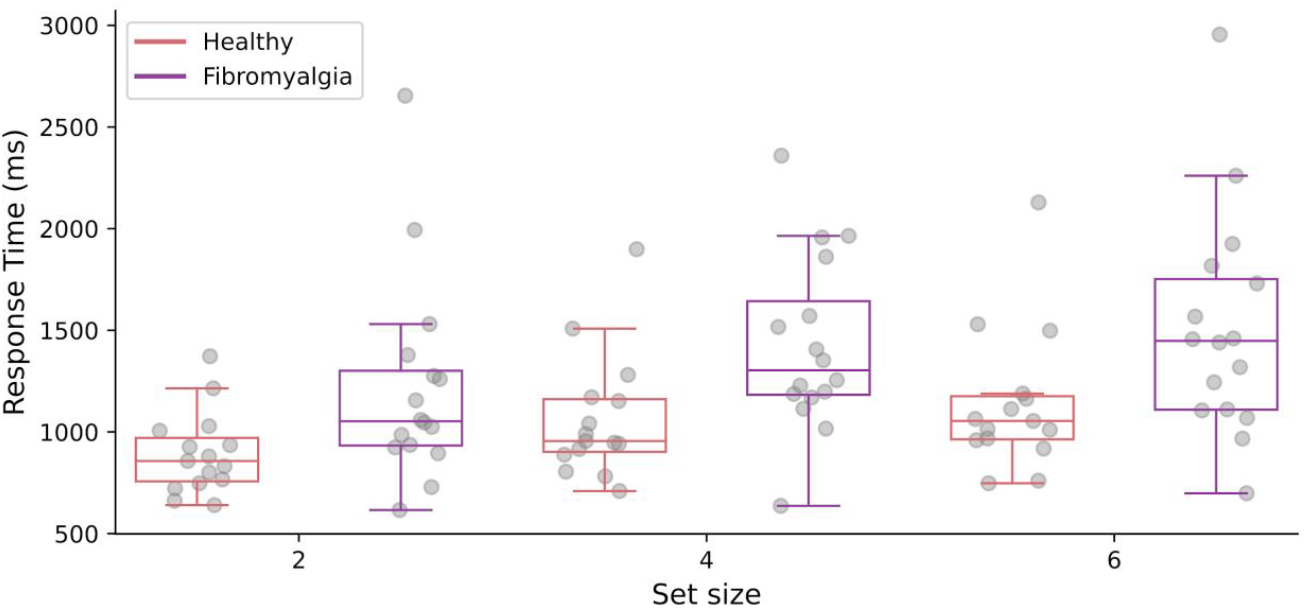
Response time for fibromyalgia patients and healthy volunteers in the Change Detection Task, for each set size

#### 3.2.2. N-back task

Figure 6 shows the distribution of the hit rate and the false alarm rate. The hit rate for fibromyalgia patients was 86.7 [27.4] % and 92.4 [10.5] % for controls. We performed a non-parametric Mann-Whitney test, and we found no statistically significant differences between the two groups (U = 110, p = 0.706). The false alarm rate for fibromyalgia patients was 9.49 [14.1] % and 5.64 [4.87] % for healthy volunteers. No significant differences were found when performing Mann-Whitney test (U = 101, p = 0.464). The response times for both groups are depicted in Figure 7. For fibromyalgia patients, the median response time was 594 [104] ms, and it was 561 [174] ms for healthy volunteers. No significant differences were found between conditions (U = 105, p = 0.572).

**Figure 6:**
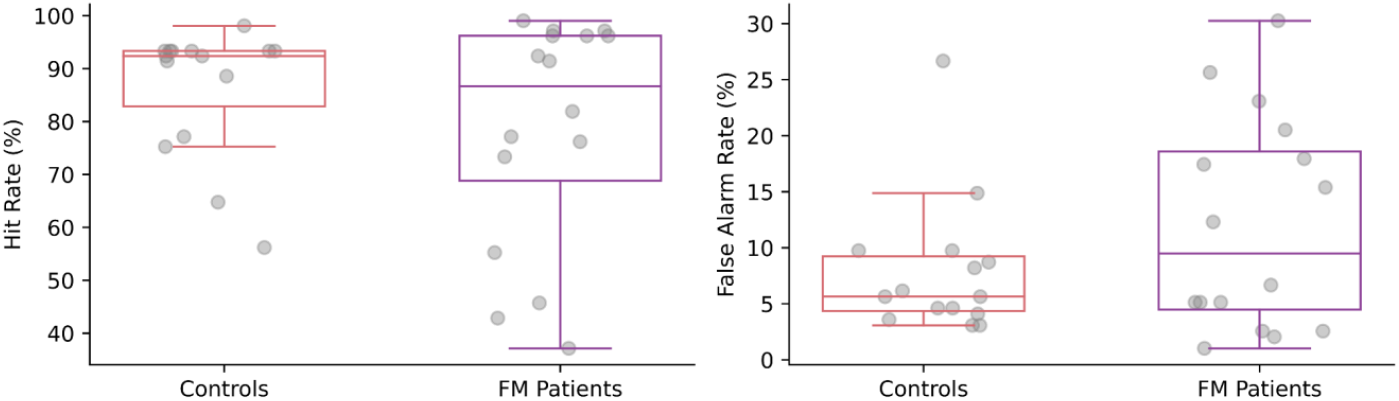
Hit rate (left) and false alarm rate (right) for fibromyalgia patients and healthy volunteers in the n-back task

**Figure 7:**
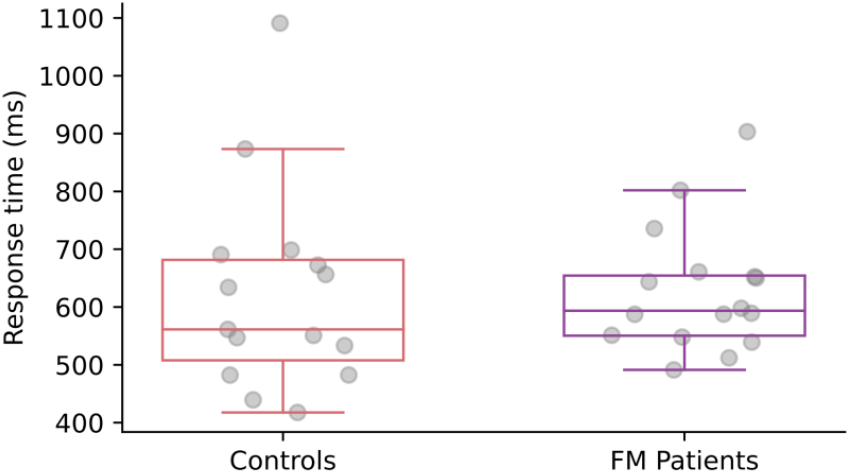
Response time for fibromyalgia patients and healthy volunteers in the n-back task

### 3.3. Electrophysiological data

EEG data from two patients and one healthy volunteer were discarded from the analysis due to substantial noise in multiple channels and blocks.

#### 3.3.1. Contralateral Delay Activity

Figure 8 shows the CDA elicited by different set sizes in fibromyalgia patients (top) and in the healthy volunteers (bottom). The negativity of the CDA is marked by the beginning of the retention interval, at approximately 0.25 s. For healthy volunteers, the greatest negativity was observed for a set size of 6, which corresponds to the higher cognitive load. The lowest negativity was observed for set size 2, corresponding to the lowest cognitive load. This behavior was not observed in the waveforms from fibromyalgia patients, although the negativity was more pronounced for set size 4 and 6. Upon statistical analysis, no significant differences were found between healthy volunteers and fibromyalgia patients for any of the cognitive loads assessed.

**Figure 8:**
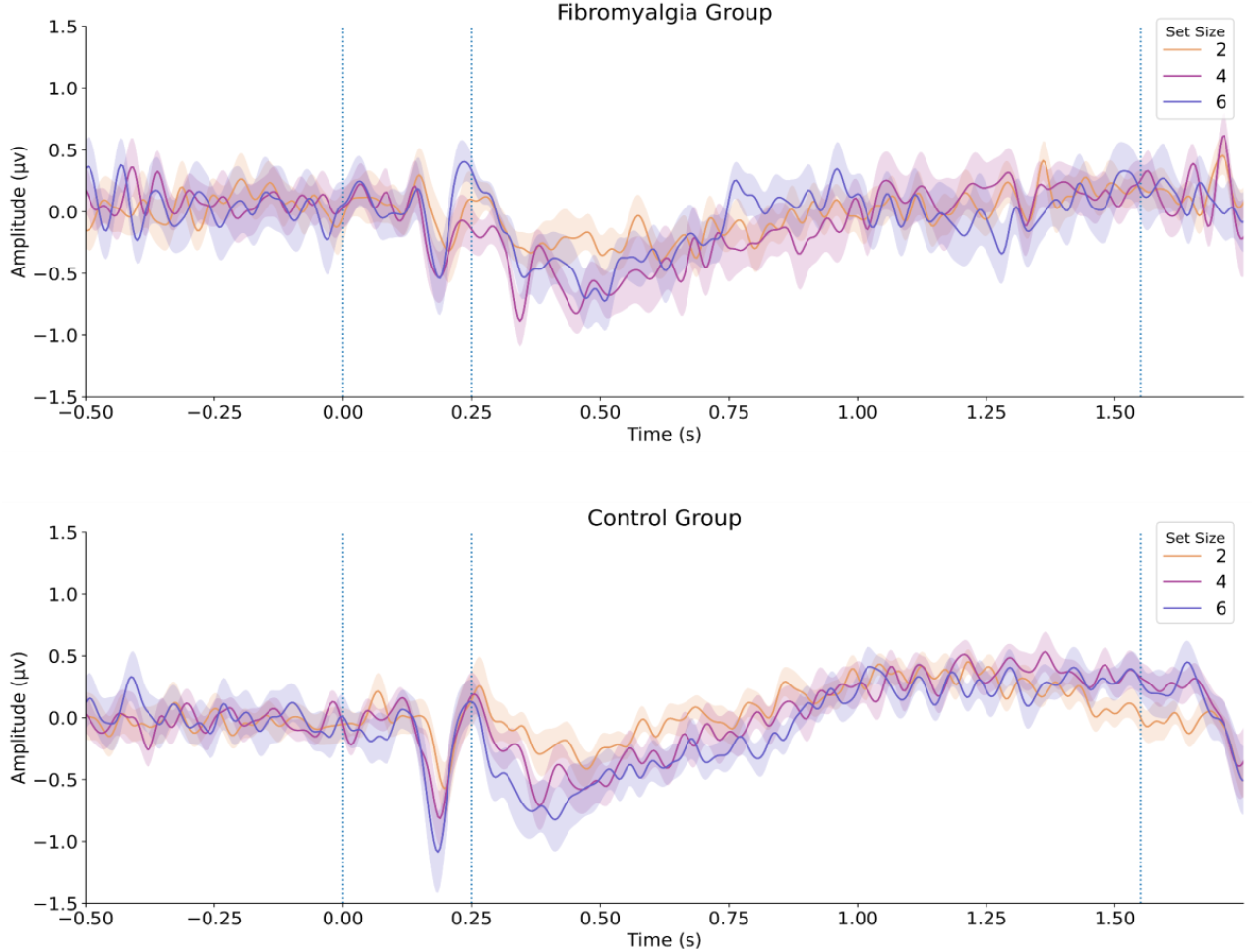
Contralateral Delay Activity for fibromyalgia patients (top) and healthy volunteers (bottom) for the different set sizes.

#### 3.3.2. N-back

Figure 9 shows the ERPs waveforms for both groups corresponding to the target (top) and non-target (bottom) stimuli at parietal location. For both groups we can observe at around 200 ms a peak consistent with P2, as well as a peak consistent with P3 at about 500 ms, more prominent among the healthy volunteers. However, no statistically significant differences were found between fibromyalgia patients and healthy volunteers for target and non-target stimuli.

**Figure 9:**
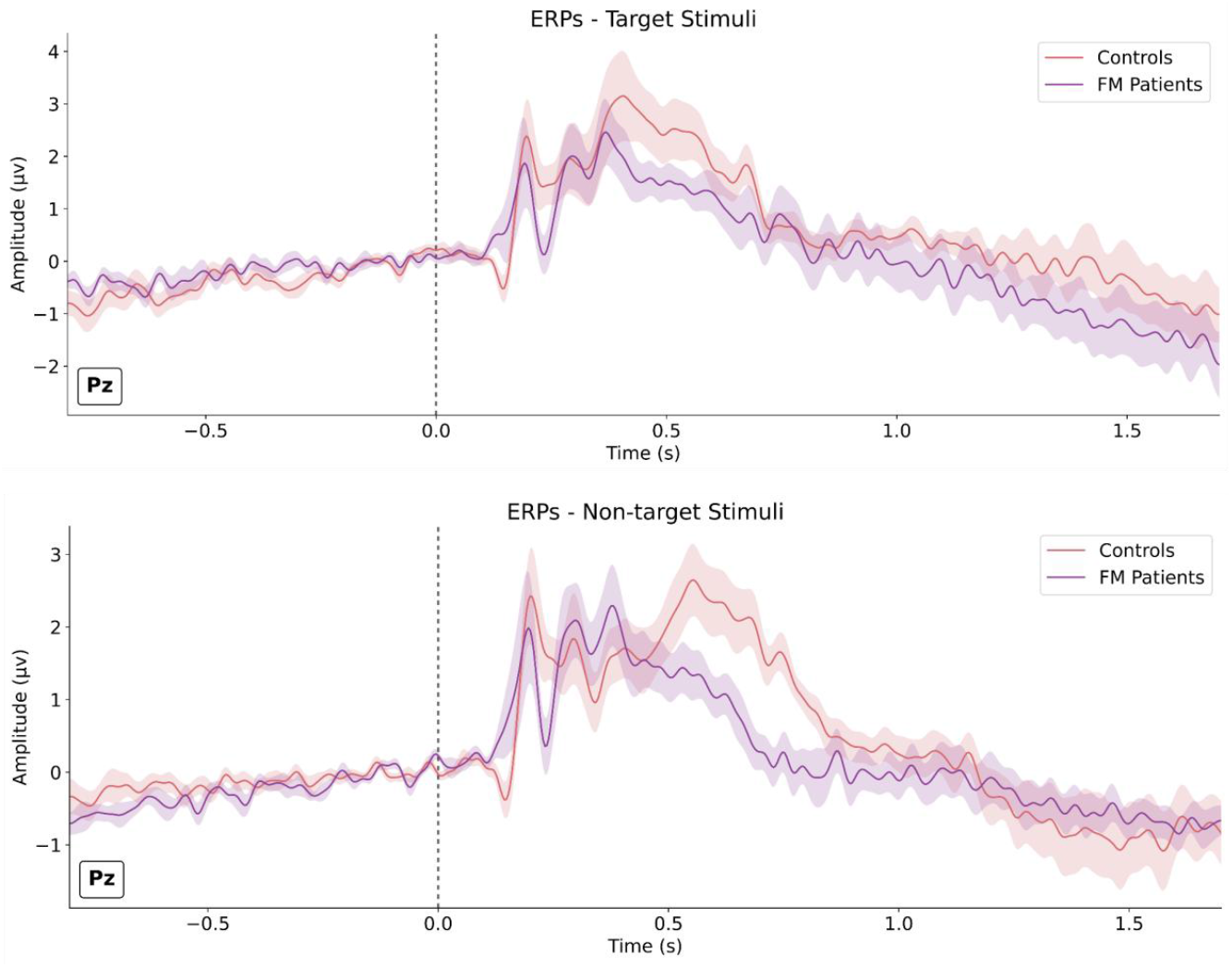
Evoked potentials for target (top) and non-target stimuli (bottom) at Pz location for fibromyalgia patients (red) and healthy participants (purple).

## 4. Discussion

Cognitive impairment, particularly related to memory deficits, is often reported by fibromyalgia patients. Several studies have focused on objective measures of these deficits, but results are inconclusive. In this study, we focused on tests designed to assess VWM, such as the change detection paradigm and the n-back task. Electrophysiological and behavioral data were simultaneously collected in a cohort of fibromyalgia patients and healthy controls.

### Evaluation of visual working memory using a change detection paradigm

Change detection paradigms have been used over the last decade as a measure of an individual’s memory capacity. Behaviorally, a quantitative estimate (K) can be derived as a combination of the hit rate and the false alarm rate. Previous studies on healthy subjects have found K to be approximately 4 (Cowan, 2001). To the best of our knowledge, no studies have focused on assessing this paradigm on a population of patients with fibromyalgia. We found that patients displayed K estimates significantly lower than those observed in healthy volunteers (1.90 [1.08] vs. 2.64 [1.55], respectively). We also observed that RT increased with higher cognitive loads. However, we did not observe a statistically significant difference, likely due to lack of sufficient power for this specific outcome. Electrophysiologically, the CDA can be observed from parieto/occipital electrodes when performing a change detection paradigm. In this study, we observed a negative deflection in the EEG recordings, consistent with CDA. However, we did not find statistically significant differences between groups or cognitive loads. The lack of group differences might be due to the fact that only correct responses are used to calculate the CDA, suggesting that the group differences might be in the number of correct responses per se, rather than in the CDA. It is also important to note that although we identified a waveform consistent with the CDA, the number of trials in this study (10 blocks of 30 trials each) was lower than typically used (21 blocks of 30 trials each (Adam et al., 2018) or 30 blocks of 52 trials each (Feldmann‐Wüstefeld, 2021)). This limitation was due to constraints in the time available for testing the patients.

### Evaluation of Visual Working Memory using a Change Detection paradigm

The n-back task has been used for the assessment of the VWM. In particular, ERP components derived from this task have been associated with processes of information encoding, as well as manipulation and updating such information (Brouwer et al., 2012; Shalchy et al., 2020). Mercado et al. (2022) focused their attention on assessing differences in ERPs between fibromyalgia patients and healthy control, finding lower amplitudes in P2 and P3 components, alongside with lower behavioral performance. In our study, such findings were not replicated, as we did not find significant differences neither in the behavioral, nor in the electrophysiological outcomes.

### Limitations and future perspectives

Several limitations should be addressed regarding our study. First, our lack of significant findings in some of the selected outcomes (particularly RT) might be due to lack of statistical power, since the sample size calculation was performed considering only the main outcome. Additional variables related to electrophysiological measures, particularly ERPs, have higher variability than behavioral outcomes, since the signal-to-noise ratio is lower, even as an average waveform (Boudewyn et al., 2018). Furthermore, the number of trials used is lower than in other studies due to time constraints. We considered that it was not ethically acceptable to have participants with fibromyalgia seated for an extended time performing tasks on a computer. In line with this, 62% of the patients that participated in the study reported a difficulty of 5 or more (in a scale from 0 to 10) to remain seated for longer than 45 minutes (See Supplementary Section). However, it cannot be ruled out that there are no sizable differences between groups. This could be explained by the phenomenon known as “*rising to the occasion*”, which involves an increase in the short-term effort to achieve a goal or complete a task that could not be maintained in the long-term (Ambrose et al., 2012). If this is the case, future work should attempt to measure the effort required to complete cognitive tasks (in an objective and/or subjective manner) and conduct a longitudinal evaluation across a longer time span. Finally, we did not ask fibromyalgia patients to withdraw or limit medication intake. Some medications prescribed for fibromyalgia can affect cognitive function, introducing a confound factor to the study. On the other hand, sudden changes in medication intake can also alter cognitive function.

## Data Availability

All data produced in the present study are available upon reasonable request to the authors

## Notes

### Competing Interest Statement

The authors have declared no competing interest.

### Clinical Protocols

https://clinicaltrials.gov/study/NCT05910372?term=Tools%20for%20the%20Differential%20Diagnosis%20of%20Fibromyalgia%20Based%20on%20Cognitive%20Tasks&rank=1

### Funding Statement

This study was partially founded by a Research and Development Project given by National University of Entre Rios.

### Author Declarations

The Central Bioethics Committee in Medical Research and Practice of Entre Rios Ministry of Health gave ethical approval for this work.

## References

Adam, K. C. S., Robison, M. K., & Vogel, E. K. (2018). Contralateral Delay Activity Tracks Fluctuations in Working Memory Performance. Journal of Cognitive Neuroscience, 30(9), 1229–1240. 10.1162/jocn_a_01233

Bell, T., Trost, Z., Buelow, M. T., Clay, O., Younger, J., Moore, D., & Crowe, M. (2018). Meta-analysis of cognitive performance in fibromyalgia. Journal of Clinical and Experimental Neuropsychology, 40(7), 698–714. 10.1080/13803395.2017.1422699

Bennett, R. M., Friend, R., Jones, K. D., Ward, R., Han, B. K., & Ross, R. L. (2009). The Revised Fibromyalgia Impact Questionnaire (FIQR): Validation and psychometric properties. Arthritis Research & Therapy, 11(4), R120. 10.1186/ar2783

Boudewyn, M. A., Luck, S. J., Farrens, J. L., & Kappenman, E. S. (2018). How many trials does it take to get a significant ERP effect? It depends. Psychophysiology, 55(6), e13049. 10.1111/psyp.13049

Brouwer, A.-M., Hogervorst, M. A., van Erp, J. B. F., Heffelaar, T., Zimmerman, P. H., & Oostenveld, R. (2012). Estimating workload using EEG spectral power and ERPs in the n-back task. Journal of Neural Engineering, 9(4), 045008. 10.1088/1741-2560/9/4/045008

Cabo-Meseguer, A., Cerdá-Olmedo, G., & Trillo-Mata, J. L. (2017). Fibromialgia: Prevalencia, perfiles epidemiológicos y costes económicos. Medicina Clínica, 149(10), 441–448. 10.1016/j.medcli.2017.06.008

Copay, A. G., Subach, B. R., Glassman, S. D., Polly, D. W., & Schuler, T. C. (2007). Understanding the minimum clinically important difference: A review of concepts and methods. The Spine Journal, 7(5), 541–546. 10.1016/j.spinee.2007.01.008

Cowan, N. (2001). The magical number 4 in short-term memory: A reconsideration of mental storage capacity. Behavioral and Brain Sciences, 24(1), 87–114. 10.1017/S0140525X01003922

Dass, R., Kalia, M., Harris, J., & Packham, T. (2023). Understanding the Experience and Impacts of Brain Fog in Chronic Pain: A Scoping Review. Canadian Journal of Pain, 7(1), 2217865. 10.1080/24740527.2023.2217865

Dizner-Golab, A., Lisowska, B., & Kosson, D. (2023). Fibromyalgia – etiology, diagnosis and treatment including perioperative management in patients with fibromyalgia. Rheumatology, 61(2), 137–148. 10.5114/reum/163094

Farina, F. R., Pragulbickaitė, G., Bennett, M., Judd, C., Walsh, K., Mitchell, S., O’Connell, R. G., & Whelan, R. (2020). Contralateral delay activity is not a robust marker of cognitive function in older adults at risk of mild cognitive impairment. European Journal of Neuroscience, 51(12), 2367–2375. 10.1111/ejn.14652

Feldmann‐Wüstefeld, T. (2021). Neural measures of working memory in a bilateral change detection task. Psychophysiology, 58(1), e13683. 10.1111/psyp.13683

Finnigan, S., O’Connell, R. G., Cummins, T. D. R., Broughton, M., & Robertson, I. H. (2011). ERP measures indicate both attention and working memory encoding decrements in aging. Psychophysiology, 48(5), 601–611. 10.1111/j.1469-8986.2010.01128.x

Gelonch, O., Garolera, M., Valls, J., Rosselló, L., & Pifarré, J. (2016). Executive function in fibromyalgia: Comparing subjective and objective measures. Comprehensive Psychiatry, 66, 113–122. 10.1016/j.comppsych.2016.01.002

Glass, J. M. (2009). Review of Cognitive Dysfunction in Fibromyalgia: A Convergence on Working Memory and Attentional Control Impairments. Rheumatic Disease Clinics of North America, 35(2), 299–311. 10.1016/j.rdc.2009.06.002

Gramfort, A. (2013). MEG and EEG data analysis with MNE-Python. Frontiers in Neuroscience, 7. 10.3389/fnins.2013.00267

Kravitz, H. M., & Katz, R. S. (2015). Fibrofog and fibromyalgia: A narrative review and implications for clinical practice. Rheumatology International, 35(7), 1115–1125. 10.1007/s00296-014-3208-7

Larson, E., Gramfort, A., Engemann, D., & Leppakangas, J. (n.d.). MNE-Python [Python]. https://zenodo.org/records/10999175

Luria, R., Balaban, H., Awh, E., & Vogel, E. K. (2016). The contralateral delay activity as a neural measure of visual working memory. Neuroscience & Biobehavioral Reviews, 62, 100–108. 10.1016/j.neubiorev.2016.01.003

Mercado, F., Ferrera, D., Fernandes-Magalhaes, R., Peláez, I., & Barjola, P. (2022). Altered Subprocesses of Working Memory in Patients with Fibromyalgia: An Event-Related Potential Study Using N -Back Task. Pain Medicine, 23(3), 475–487. 10.1093/pm/pnab190

Mista, C. A., Intelangelo, L., & Biurrun Manresa, J. (2023). Personalized pain management: The relationship between clinical relevance and reliability of measurements. European Journal of Pain, 27(9), 1056–1064. 10.1002/ejp.2110

Pidal-Miranda, M., González-Villar, A. J., Carrillo-de-la-Peña, M. T., Andrade, E., & Rodríguez-Salgado, D. (2018). Broad cognitive complaints but subtle objective working memory impairment in fibromyalgia patients. PeerJ, 6, e5907. 10.7717/peerj.5907

Rouder, J. N., Morey, R. D., Morey, C. C., & Cowan, N. (2011). How to measure working memory capacity in the change detection paradigm. Psychonomic Bulletin & Review, 18(2), 324–330. 10.3758/s13423-011-0055-3

Salgueiro, M., García-Leiva, J. M., Ballesteros, J., Hidalgo, J., Molina, R., & Calandre, E. P. (2013). Validation of a Spanish version of the Revised Fibromyalgia Impact Questionnaire (FIQR). Health and Quality of Life Outcomes, 11(1), 132. 10.1186/1477-7525-11-132

Schober, P., Bossers, S. M., & Schwarte, L. A. (2018). Statistical Significance Versus Clinical Importance of Observed Effect Sizes: What Do P Values and Confidence Intervals Really Represent? Anesthesia & Analgesia, 126(3), 1068–1072. 10.1213/ANE.0000000000002798

Shalchy, M. A., Pergher, V., Pahor, A., Van Hulle, M. M., & Seitz, A. R. (2020). N-Back Related ERPs Depend on Stimulus Type, Task Structure, Pre-processing, and Lab Factors. Frontiers in Human Neuroscience, 14, 549966. 10.3389/fnhum.2020.549966

Sullivan, G. M., & Feinn, R. (2012). Using Effect Size—Or Why the P Value Is Not Enough. Journal of Graduate Medical Education, 4(3), 279–282. 10.4300/JGME-D-12-00156.1

Tesio, V., Torta, D. M. E., Colonna, F., Leombruni, P., Ghiggia, A., Fusaro, E., Geminiani, G. C., Torta, R., & Castelli, L. (2015). Are Fibromyalgia Patients Cognitively Impaired? Objective and Subjective Neuropsychological Evidence. Arthritis Care & Research, 67(1), 143–150. 10.1002/acr.22403

Vallejo, M. A., Rivera, J., Esteve-Vives, J., & Rodríguez-Muñoz, M. F. (2012). Uso del cuestionario Hospital Anxiety and Depression Scale (HADS) para evaluar la ansiedad y la depresión en pacientes con fibromialgia. Revista de Psiquiatría y Salud Mental, 5(2), 107–114. 10.1016/j.rpsm.2012.01.003

Walitt, B., Čeko, M., Khatiwada, M., Gracely, J. L., Rayhan, R., VanMeter, J. W., & Gracely, R. H. (2016). Characterizing “fibrofog”: Subjective appraisal, objective performance, and task-related brain activity during a working memory task. NeuroImage: Clinical, 11, 173–180. 10.1016/j.nicl.2016.01.021

Wiegand, I., Hennig-Fast, K., Kilian, B., Müller, H. J., Töllner, T., Möller, H.-J., Engel, R. R., & Finke, K. (2016). EEG correlates of visual short-term memory as neuro-cognitive endophenotypes of ADHD. Neuropsychologia, 85, 91–99. 10.1016/j.neuropsychologia.2016.03.011

Williams, D. A., & Arnold, L. M. (2011). Measures of fibromyalgia: Fibromyalgia Impact Questionnaire (FIQ), Brief Pain Inventory (BPI), Multidimensional Fatigue Inventory (MFI-20), Medical Outcomes Study (MOS) Sleep Scale, and Multiple Ability Self-Report Questionnaire (MASQ). Arthritis Care & Research, 63(S11), S86–S97. 10.1002/acr.20531

Wolfe, F., Clauw, D. J., Fitzcharles, M.-A., Goldenberg, D. L., Katz, R. S., Mease, P., Russell, A. S., Russell, I. J., Winfield, J. B., & Yunus, M. B. (2010). The American College of Rheumatology Preliminary Diagnostic Criteria for Fibromyalgia and Measurement of Symptom Severity. Arthritis Care & Research, 62(5), 600–610. 10.1002/acr.20140

